# Automated multi-scale computational pathotyping (AMSCP) of inflamed synovial tissue

**DOI:** 10.1101/2023.05.21.23290242

**Authors:** Richard D. Bell, Matthew Brendel, Maxwell Konnaris, Justin Xiang, Miguel Otero, Mark A. Fontana, Accelerating Medicines Partnership Rheumatoid Arthritis and Systemic Lupus Erythematosus (AMP RA/SLE) Consortium, Edward DiCarlo, Jennifer Anolik, Laura Donlin, Dana Orange, H. Mark Kenney, Edward M. Schwarz, Lionel B Ivashkiv, Fei Wang

**Author notes:** Authors contributed equally (Richard D. Bell, Matthew Brendel, Edward M. Schwarz, Lionel B Ivashkiv, Fei Wang). Corresponding Author: Richard D. Bell, PhD 515 E 71^st^ New York, NY.

## Abstract

Rheumatoid arthritis (RA) is a complex immune-mediated inflammatory disorder in which patients suffer from inflammatory-erosive arthritis. Recent advances on histopathology heterogeneity of RA pannus tissue revealed three distinct phenotypes based on cellular composition (pauci-immune, diffuse and lymphoid), suggesting distinct etiologies that warrant specific targeted therapy. Thus, cost-effective alternatives to clinical pathology phenotyping are needed for research and disparate healthcare. To this end, we developed an automated multi-scale computational pathotyping (AMSCP) pipeline with two distinct components that can be leveraged together or independently: 1) segmentation of different tissue types to characterize tissue-level changes, and 2) cell type classification within each tissue compartment that assesses change across disease states. Initial training and validation were completed on 264 knee histology sections from mice with TNF-transgenic (n=233) and injected zymosan induced (n=32) inflammatory arthritis. Peak tissue segmentation performance with a frequency weighted mean intersection over union was 0.94 ± 0.01 and peak cell classification F1 was 0.83 ± 0.12.

We then leveraged these models and adapted them to analyze RA pannus tissue clinically phenotyped as pauci-immune (n=5), diffuse (n=28) and lymphoid (n=27), achieving peak cell classification performance with F1 score of 0.81 ± 0.06. Regression analysis demonstrated a highly significant correlation between AMSCP of lymphocyte counts and average Krenn Inflammation Score (rho = 0.88; p<0.0001). While a simple threshold of 1.1% of plasma cells demonstrated the phenotyping potential of our automated approach vs. a clinical pathologist with a sensitivity and specificity of 0.81 and 0.73. Taken together, we find AMSCP to be a valuable cost-effective method for research. Follow-up studies to assess its clinical utility are warranted.

## Introduction

Disease pathotyping with histopathology, the discovery of disease subtypes using target organ histology, is a critical step in understanding etiology and response to therapy in heterogeneous diseases, like rheumatoid arthritis (RA). Our understanding of RA, which is a chronic, inflammatory joint disease, has greatly benefited from histopathology subtyping because the disease has distinct and disparate etiologies with largely stable pathotypes [1, 2] that show differential response to therapy [3–7]. However, the process of pathotyping a patient can be resource intensive involving both basic and immune-stains requiring a high level of expertise by pathologists to interpret tissue and cellular histologic features, and prone to inter- and intra-observer variations [8, 9]. More cost effective and efficient procedures need to be developed in order incorporate these types of data into a precision medicine decision making process.

Recent work describing RA pathotypes uncover three distinct synovial pathotypes 1) cellular dense, lymphocyte rich (lymphoid), 2) myeloid rich with few lymphocytes (diffuse/myeloid), and 3) fibroblast rich (pauci-immune); which are identifiable through distinct cellular and tissue level changes within synovial joint biopsies [10–14]. These pathotypes also correlate with antibody positivity (i.e. anti-citrullinated peptide antibodies) with the lymphoid type enriched in antibody positive patients whereas both the diffuse/myeloid and pauci-immune types have equal contributions of both antibody positive and negative patients. This aligns with preclinical models that rely on antibody dependent (e.g. Collagen Induced Arthritis and Serum Induced Arthritis) and independent mechanism (e.g. humanized TNF Transgenic and Zymosan Induced Arthritis) to study disease and are phenotypically similar to these pathotypes [15]. Thue, tools that allow us to study both murine and human pathology would improve our overall understanding of this heterogeneous disease.

Computational tools to study histological changes have been shown to augment pathologist workflows and allow for the identification of disease specific patterns [16]. In particular, machine learning, and specifically deep learning, is a data-driven framework that has recently had success in the automated analysis of musculoskeletal imaging data [17].

Additionally, computational tools can automate and provide a more holistic analysis a wide variety of histopathologic tissue and cell-level changes to enable a more detailed understanding of disease subtypes. However, automated and comprehensive tool to study both tissue and cell type specific changes in arthritis that can quantify therapeutic or clinically meaningful differences has not yet been developed [17].

In this work, we developed an automated multi-scale computational pathotyping (AMSCP) model to analyze tissue and cell-level changes during the progression of inflammatory arthritis. This model can pathotype both mouse and human inflammatory arthritis in therapeutic intervention studies and clinical meaningful scenarios. We leveraged innovative transfer- and active-learning techniques to improve model performance and workload efficiency. Our modeling framework consists of two distinct components that can be utilized together or independently, (1) a method to segment different tissue types to characterize tissue-level changes and (2) a method to classify cell types within each tissue compartment to study how these change across disease states. We utilized two mouse models of inflammatory arthritis to train and validate our computational models with subsequent implementation on additional datasets to measure therapeutic efficacy, known biologic differences and discover novel pathologic changes. Then, we utilize a human synovial biopsy data set from the Accelerating Medicines Partnership Rheumatoid Arthritis (AMP-RA) to demonstrate our model’s utility in a clinical setting by classifying lymphoid pathotypes from diffuse/myeloid pathotype and identifying cell types associated with the pauci-immune pathotype while preserving spatial cell-level data.

## Methods

### Dataset Description

All mouse work was approve by the University Committee on Animal Resources at the University of Rochester Medical Center and the Institutional Animal Care and Use Committee at the Hospital for Special Surgery. Whole slide images (WSIs) of sagittal mouse knee sections were taken from two different mouse models of inflammatory arthritis and the accompanying controls for segmentation experiments (**Supplemental Figure 1A**). Batch A consisted of male and female TNF Transgenic mice (TNF-Tg, n=47) and wild-type littermates (WT, n = 15) used in a previous publication [18, 19]. Batch B consisted of male and female knees that received intra-articular injections of 180ug of Zymosan to induce Zymosan Induced Arthritis (ZIA, n = 24) and control contralateral limbs (Control, n = 8) [20] that were euthanized on Day 7 after injection. Different batches were used to test model generalizability across different biological mechanisms of arthritis development, differences in H&E staining protocols and slide scanner used to digitally capture slides at 40x magnification (Batch A: VS120 Olympus, 0.173 μm per pixel; Batch B: CS2 Aperio Leica, 0.253 μm per pixel).

To further test model generalizability, 2 different holdout datasets were used to validate the model: 1) H&E-stained sagittal knee WSIs from Bell et al [18, 19] and 2) Orange G-H&E stained sagittal knee WSIs from Kenney et al [21]. Slides from Bell et al were ensured to not be used in the initial model training, internal validation or testing. These included slides from 6-month-old male TNF-Tg (n=33 slides from 14 knees) and WT littermates (n=43 slides from 17 knees), and slides from 9.5 month old male TNF-Tg mice (Anti-TNF: n=24 slides from 8 knees; Placebo: n= 29 slides from 10 knees) and WT littermates (Placebo: n=42 slides from 15 knees) either treated with a 6 week course of Anti-TNF antibodies or Placebo control. To generate downstream measurements of tissues of interest, a region of interest (ROI) was drawn from the tibial growth plate to femoral growth plate including the anterior and posterior extra articular tissue.

WSIs of H&E-stained human synovial biopsies were collected from the Accelerating Medicines Partnership Rheumatoid Arthritis (AMP-RA) Phase II consortium [22]. In short, synovial tissue biopsies were acquired from RA patients at 13 different clinical sites in the United States and 2 from the United Kingdom from October 2016 to February 2020. The study was performed in accordance with protocols approved by the institutional review board at each site. The tissue was paraffin embedded, stained with H&E and imaged on a VS120 Olympus. Board certified pathologists in the AMP-RA consortium classified each case as either lymphoid (n = 27), diffuse/myeloid (n=28) or pauci-immune (n=5), and provided the Krenn inflammation score for each case [23].

### Semantic Segmentation Annotation and Preprocessing

Manual annotations were performed within QuPath [24] to assign labels for WSIs. To test model performance across tissue types at different granularity, multiple different class structures were tested (**Supplemental Figure 2**). Eleven different classes were manually annotated, including synovium, muscle and tendon, growth plate, bone marrow, cortical bone, trabecular bone, articular cartilage, meniscus, fat, bone marrow fat, and histology artifact (i.e., out of focus). A seven-class, nine-class, and ten-class segmentation task was generated by merging histologically similar tissues, such as merging cartilage and meniscus into the same class (**Supplemental Figure 2**). Overall, we estimate about 250 hours were spent annotating the 11 tissue classes on 94 WSIs.

Due to the gigapixel nature of WSIs, the entire slide cannot be fed directly into a deep learning model. Previous work has shown that WSIs can be broken into patches, in this case 512 pixel x 512 pixel, to perform downstream learning tasks [25]. For semantic segmentation, a custom QuPath script was used to export patches at a 4x downsample while filtering out regions of the scanned slide that lacked annotations or without tissue. Additionally, images were normalized to mean 0 and standard deviation 1 by sampling a subset of patches to get mean and standard deviation RGB statistics.

### Semantic Segmentation Models and Training Strategy

Two different training strategies were employed to test model performances (**Supplemental Figure 1B**). First, a mixed training strategy was used, in which slides from Batch A and Batch B were mixed and randomly assigned to a training, validation, and test split, using 70%, 15%, and 15% for each split, respectively (3 splits total). In this training strategy, stratification was performed by batch and disease type to ensure even splitting into each set, and each patch was split on the slide level to prevent data leakage across splits. Next a single batch training strategy was tested to assess how site-specific differences in histology slides may impact model performance [26]. In this training strategy, Batch A was used for the training and validation sets, and Batch B was used as the held-out test set.

Data augmentation strategies were also tested to assess their impact across the different training strategies (**Supplemental Figure 3**). We had three different levels of augmentation tested, (1) None, (2) Low, (3) Medium, and (4) High. The python package imgaug[27] as used to implement augmentation. Augmentation was applied in the following way: (1) None had zero augmentation, (2) Low had 2-4 augmentation process applied 25% of the time, (3) Medium had 3-7 augmentations applied 50% of the time and (4) High augmentation had 5-11 augmentation process applied 50% of the time during training. Augmentation was randomly selected from 11 different types of augmentations including, horizontal flip (p = 0.5), coarse dropout or pixel dropout from 0.2x the original image resolution (p=0.1), one of three different rotation types at 90°, 180°, and 270°, additive gaussian noise sampled from a normal distribution with mean 0 and variance 0.2*255, blur using gaussian kernel with sigma of 1.5, hue modification using addition (-30,10) and saturation modification using multiplication (0.5,1.5) and linear contrast (0.5,2), brightness adjustment both add(-30,30) and multiply (0.5,1.5), and color change adjustment (3000,8000).

To compare segmentation performance using both conventional machine learning and deep learning methods, we tested two different model architectures. A Random Forest (RF) model implemented with OpenCV [28] in QuPath and an U-Net++ [29] deep learning model implemented in PyTorch [30]. To train the RF, we first applied a Simple Linear Iterative Clustering algorithm (σ = 5, spacing = 20 μm, Max Iterations = 1, Regularization = 0.01) in which each over segmented area was considered a super-pixel. We then performed color and shape feature extraction of each super-pixel, and calculated the average features of super-pixels within 40 and 80 μms. These features were then used in the RF and the tissue class of each super-pixel was used to supervise the model. After the RF was trained, its performance was assessed fully in QuPath. For our deep learning pipeline, we utilized the segmentation_models_pytorch [31] package for the UNET++ implementation with an ImageNet pre-trained EfficientNet-B5 backbone for the encoder [32]. We utilized the combo loss, which was the arithmetic average of the dice loss and binary cross entropy (BCE) loss during model training [33]. A learning rate scheduler was used to train the model using a step size of 2 and a gamma of 0.5. The learning rate started at 0.025 using the stochastic gradient descent (SGD) optimizer with momentum (0.9) and regularization of 1x10^-4^ and was trained for 10 epochs.

When jointly training across 10 different classes, this model provided no prediction for the meniscus class resulting in a mIOU of 0 ± 0 (Supplemental Figure 6), likely due to insufficient training examples (Supplemental Figure 2; 0.9% frequency overall). As the meniscus was important for downstream biological analyses, we developed a strategy to improve predictions for this class. A second U-Net++ model was fine-tuned from the nine-class model (i.e. the model that has cartilage and meniscus merged into one class) by changing the prediction (final) layer specifically to predict between cartilage and meniscus. We then re-trained the full model only on images and Ground Truth annotations (GTs) that contained either cartilage or meniscus within the mixed training set.

Previous work had suggested that U-Net based semantic segmentation can have image boundary level artifacts [34]. Therefore, we assessed how including patch level overlap for prediction can improve model performance. We included experiments using no overlap, 50% overlap and 66% overlap between them with images from both batches (**Supplemental Figure 4**). To analyze the results, we loop through all the predictions (*N*) for the entire WSI and calculate a majority vote for each pixel after thresholding to removes low confidence predictions (pixel value of 75). *N* can be variable depending upon the region, for example if it is on border, but typically is between 4-9.

### Semantic Segmentation Model Evaluation, Inference on External Validation Set and Statistical Evaluation

To evaluate the semantic segmentation model performance for the validation and test set slides with ground-truth labels, we calculated the mean Intersection over Union (mIOU) and frequency-weighted mIOU to prevent very rare classes from drastically impacting overall model performance [35]. All model hyperparameter optimization was performed on the validation set, and once the above parameters were chosen the models were used to infer segmentation on the test set and the mIOUs were calculated.

We used the optimal settings from the training/validation process to evaluate the model performance on the held-out datasets. Pearson’s correlation was used to compare the hand drawn synovial tissue area reported in Bell et al [18] with model classified synovial tissue area. The fine-tuned 10-class model was used to infer tissue segmentation on the held-out data [21, 36]. Specifically, the UNET++ 9 class model was used to predict tissues classes on each patch, and if the nine-class model had a prediction output for the combined cartilage and meniscus class on a patch, then the patch was passed through the fine-tuned 2 class model to assign to either the meniscus or cartilage class. Predictions were merged by only allowing the fined tuned predictions to be within the predictions from the combined Cartilage-Meniscus class. Once the inference was complete, a ROI was drawn on each slide from femoral growth plate to tibial growth plate around the joint to restrict the downstream analysis to the joint space, subchondral bone, and synovial adjacent tissue. Tissue area was calculated and a One-Way ANOVA with Tukey’s post-hoc adjustment was used to detect significant differences.

### Cell Type Classification Framework and Preprocessing

For cell type classification, a combination of transfer learning and active learning was used to identify several different cell types that exist within the joint tissue. Cell type classification can be broken into a two-step process, (1) segmentation and (2) classification. For cell segmentation, transfer learning was used by leveraging a deep learning model, HoVer-Net [37], pretrained on the PanNuke dataset [38], to extract nuclei regions. Image patches (1024 x 1024 pixels) at 40x magnification were given as inputs, and ROI contours of nuclei were obtained to perform feature extraction upon. These nuclei with their features and labels (detailed below) were then leveraged in a gradient boosted decision tree (GBDT) model to classify cells.

The input for the classifier was derived from features extracted from each ROI generated by HoVer-Net. Specifically, every nuclei from the json file output of HoVer-Net was converted into a ROI (.roi) file to be read into FIJI/ImageJ [39] for feature extraction using a custom script. Detailed workflow for the ImageJ/FIJI analysis is described as follows. Each image was split using the built-in color deconvolution [40] algorithm in FIJI into hematoxylin and eosin color channels. For each channel the nuclei were measured for several different parameters, including morphological quantities (area, perimeter, circularity, feret’s diameter, feret angle, aspect ratio, roundness, and solidity) and staining color quantities (mean, mode, min, max, standard deviation, skewness, median, and kurtosis), and the nuclei ROI was enlarged to calculate cell specific cytoplasmic H&E color information for each cell. Characteristics of neighboring cells at several different distance ranges were used to include tissue level context into cell type classification [41]. A final total of 854 features were extracted for the downstream analysis.

Known healthy and pathologic cell types that contribute to inflammatory arthritis were then annotated on both mouse and human tissues (**Supplemental Figures 8 and 10**). The mouse proof-of-concept classification task consisted of bone-embedded cells, vessel cells, adipo-stromal cells (both adipose and stromal cells within fatty tissue), synovial fibroblasts (healthy and pathologic), chondrocytes, lymphocytes, and other synovial lining cells (healthy and pathologic) as detailed in **Supplemental Figures 8**; annotated by subject-matter experts familiar with histologic analysis of these cell types. These cells were annotated on six healthy, eight mild disease and five severely diseased TNF-Tg knee sections. For human samples, a clinically meaningful set of cell types were labeled by a senior pathologist, following a standard cell type hierarchy (**Supplemental Figure 10**). These included stromal/connective tissue cells, synovial lining cells, synovial fibroblasts, vascular endothelial cells, tissue macrophages/histocytes, lymphocytes, and plasma cells. These cells were labeled on five lymphoid, five diffuse and three pauci-immune cases. Nuclei were then mapped to manually annotated nuclei by checking if a nuclei’s centroid (as determined by Hover-Net) was within an annotation mask.

### Cell Type Classification Model and Active Learning

A total of 4,712 cells were annotated for mouse cell type classification from seven different classes. Cells were labeled from a total of 19 different slides. A GBDT model was trained for cell type classification, using 5-fold cross validation to select the best models. To evaluate model performance, cell types were predicted for two different biological settings, (1) to identify cell composition changes across different disease severities on the remaining cells (>300,000) on the 19 slides, and (2) identify differences between male and female mice in the context of disease progression in a held-out dataset [18]. Finally, average synovial inflammatory infiltrate scores and average pannus invasion scores were correlated with lymphocyte and synovial lining cell counts respectively (Spearman’s Correlation).

Human annotation was the time-consuming step for the cell type classification pipeline. Therefore, we applied the active learning strategy to improve the annotation efficiency for cell annotation of human samples. To develop this strategy, we tested a proof-of-concept active learning strategy using labeled data from the mouse H&E slides (**Supplemental Figure 9**).

Active learning is an iterative process that consists of three main steps, (1) annotation, (2) model training, and (3) sample selection for further annotations. Its goal is to select the samples that can lead to the largest model performance improvement when adding to the training data after annotation. To validate the strategy, 100 different rounds of 5-fold cross-validation were performed. Average F1-scores were reported for each class and a macro-F1 score was additionally reported. 25 runs of 5-fold cross validation were removed due to cells from a single class not being present in both the training and testing sets. For the training dataset for each split, 5% of cells were first randomly selected as the first set of cells selected as being labeled annotated. Subsequently, the GBDT classifier was trained using this randomly selected data. Several different metrics for determining cells for annotation and subsequent model finetuning, including smallest margin uncertainty [42], least confidence uncertainty [43] and entropy-based uncertainty [42] were assessed. The top 5% of cells were added to the training dataset and the cycle of model training and evaluation and new cell annotations continued until the entire training dataset was used. A random selection of cells after shuffling was also tested to compare model performance to the various active learning strategies. The package modal [44] was leveraged in our implementation. Mean and 95% confidence intervals are reports for each subset across the 75 different runs of 5-fold cross validation.

### Cell Classification Model Evaluations

Confusion matrices were generated for model prediction along with F1-scores calculated as 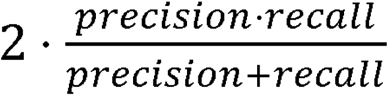, where 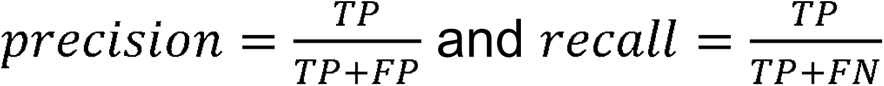, where TP, FP, FN stand for true positives, false positives, false negatives. Models were tested using known cell types within specific tissue types to evaluate the model qualitatively.

### Analysis of Human Synovial Biopsy Slides

Active learning was then leveraged for human cell type classification using H&E-stained slides of human synovial biopsy tissue of RA patients from the AMP consortium as described above. Multiple rounds of cell type labeling were performed with the assistance of active learning, to obtain a total of 2,639 cells grouped in seven different cell types, detailed in Supplemental Figure 9 (stromal/connective tissue cells n=597, synovial lining cells n= 309, synovial fibroblasts n = 189, vascular endothelial cells n = 486, Tissue Macrophages/Histocytes n = 201, lymphocytes n = 826, and plasma cells n = 310). A cell type classification model using GBDT with five-fold cross validation was trained on this dataset and inferences from the best performing model applied to all cells on the slides within this patient cohort. Summary cell type quantification (total cell counts and percent of total) was then assessed for each patient. Two analyses were performed using the derived cell types from the cell classification model. First cell type proportions were analyzed across different pathotypes. Specifically, statistical significance testing using lymphocyte, plasma cell, and fibroblast slide proportions were evaluated across pathotypes. Second, pathologist-derived, and clinically relevant Krenn inflammation scores were correlated with Spearman’s correlation to the counts of the lymphocytes for each slide to assess disease severity across human samples.

### Visualization of Data

Uniform Manifold Approximation and Projection [45] visualization was used for feature representations between batches and cell type framework features. Masks for each class were reimported into QuPath [24] for visualization purposes.

### Dataset and Code Availability

All original data and analysis scripts will be provided upon reasonable request.

## Results

### Deep Learning Segmentation Can Identify Major Tissue Types within Mouse Knee Histology and Measure Therapeutic Response

Several model training choices, including patch overlap, training strategy, and use of different amounts of augmentation during training, were empirically derived from initial experiments to inform the final training of the deep learning segmentation model (UNET++) [34, 46]. First, we tested if 0%, 50% or 66% patch overlap was more performative and determined that 66% overlap performed the best (0% Overlap fwIOU: 0.72 ± 0.04; 50% Overlap fwIOU: 0.93 ± 0.02; 66% Overlap fwIOU: 0.95 ± 0.01; **Supplemental Figure 4A**). Qualitatively, there were less tiling edge artifacts in the 66% overlap vs 50% overlap results (50% - Arrows **Supplemental Figure 4B** vs **Figure 1**). Second, a mixed training strategy was shown to overcome the large staining batch effect (**Supplemental Figure 5A**) commonly seen in histology datasets with all levels of augmentation equally performative (**Supplemental Figure 5B**). However, if the data is restricted to a single batch and data augmentation is introduced, model performance for the single batch training strategy becomes comparable to model performance using the mixed strategy (High Augmentation UNET++: 0.81± 0.02 vs 0.80 ± 0.06 mIOU for mixed and single batch respectively), demonstrating the need for augmentation to generalize across batches with this level of heterogeneity in staining and other imaging variations (**Supplemental Figure 5B and C**). Thus, we choose to employ a mixed training strategy, with 66% patch overlap and with high augmentation to optimize model performance and generalizability.

**Figure 1.**
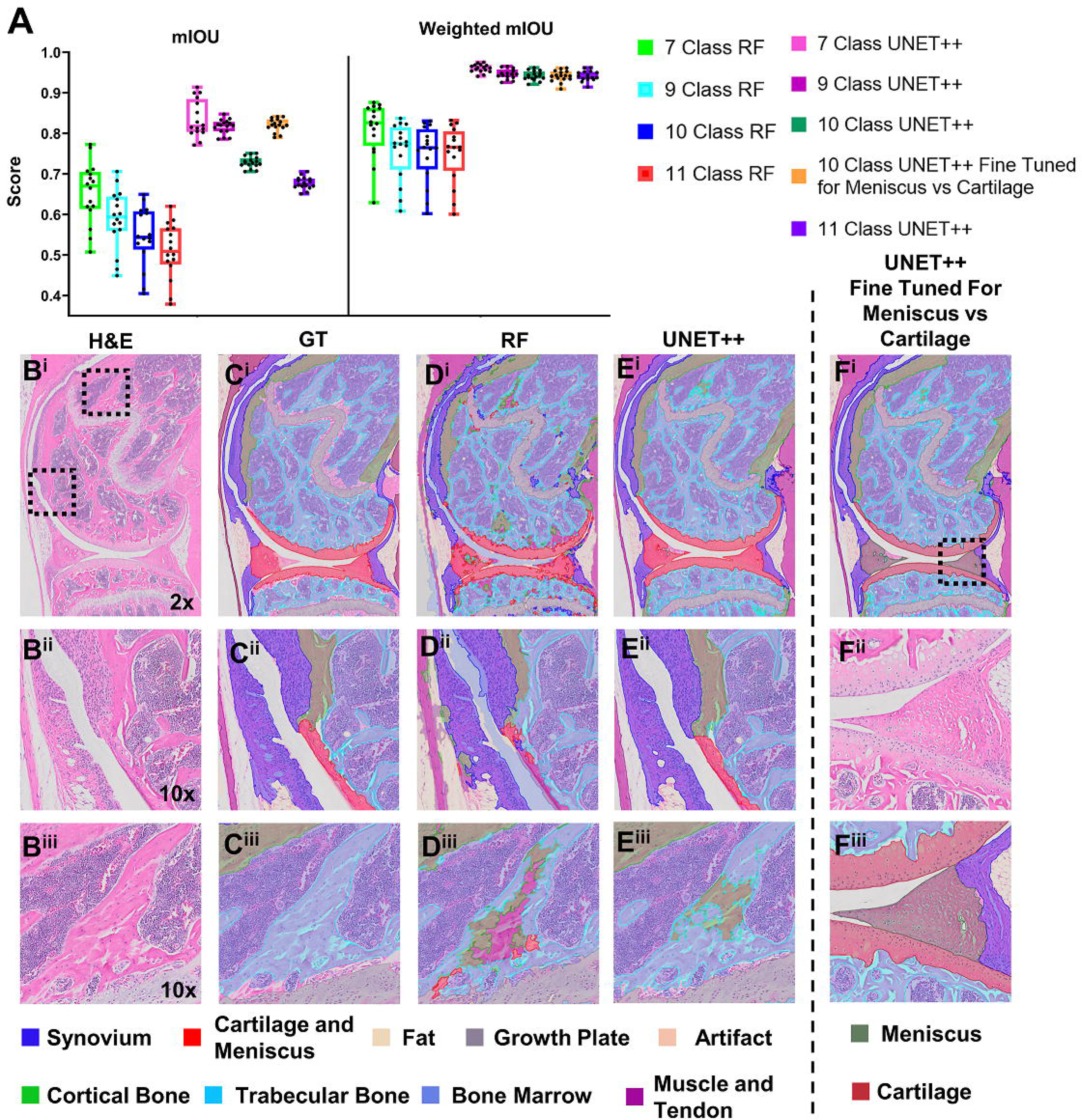
A fine-tuned 10 class model can segment relevant tissue in inflammatory arthritis. Mean Intersection over union (mIOU) and class frequency weighted mIOU statistics from the RF and DL segmentation models are presented in A, and demonstrate that UNET++ 9 Class model that was then fine tuned to segment the meniscus and cartilage performs the best. Representative images of H&E (B) image, with Ground Truth (GT, C), RF (D), UNET++ (E) and the Fine-Tuned model’s prediction overlays demonstrate the successful segmentation and mistakes. Specifically, the area where the synovial pannus tissue reaches the cartilage (B^ii^ - E^ii^), the RF model (D^ii^) underperforms while the UNET++ (E^ii^) are almost perfect. A common mistake within all models is misclassifying the trabecular bone (B^iii^ - E^iii^) for other tissues, with the RF (D^iii^) model misclasses for cartilage, muscle and tendon, and cortical bone; while UNET++ (E^iii^) misclass only as cortical bone. When investigating the overall misclassification of cortical bone for trabecular bone it occurs in <5% of all tissue area. Importantly, the Fine-Tuned model (F) successfully segments the Cartilage and Meniscus from each other (F^ii^ and F^iii^).

Once the training strategy was established, model performance was benchmarked across segmentation tasks at multiple tissue granularities and compared with a standard RF model built-in to QuPath (**Figure 1**). As expected, as the number of different tissue types increases, model performance decreases across both the RF model and UNET++ model, and the DL model outperform the RF at all levels. Interestingly, the magnitude of change is smaller for the UNET++ model compared to the RF model. When testing the UNET++ model, using the ten-class granularity, model performance drops from 0.88 ± 0.06 mIOU for the cartilage and meniscus class to 0.83 ± 0.05 mIOU for the cartilage class and 0.0 ± 0.0 mIOU for the meniscus class indicating a complete loss of meniscus identification (**Supplemental Figure 6**). Because defining the amount of cartilage and meniscus is an incredibility important pathologic readout in joint diseases (e.g. pannus invasion at end stage arthritis), we developed a finely tuned two class model and placed it sequentially after the 9-class model. Predictions from both were incorporated during majority voting process and performance jumps for this fine-tuned resulting ten-class model from 0.72 ± 0.01 mIOU to 0.82 ± 0.02 mIOU (**Figure 1A and B**). Specifically, performance for the cartilage and meniscus classes were 0.9 ± 0.04 and 0.92 ± 0.05, respectively. We additionally observe that the worst performing classes are the artifact class and bone marrow fat class, the two most infrequent classes, suggesting that fine tuning may work to improve performance for one or both (**Supplemental Figure 6)**. Thus, for all subsequent work we either used the 9-class model (termed Original model) for ease of computation or the Fine-Tuned 10 class model for meniscus segmentation because these provided the best predictive performance while encompassing the most amount of tissues.

After training the model on the mouse cohorts as described, the model was then externally validated (i.e., completely independent of the training, testing and validation process above) in two ways. 1) We first validated our Fine-Tuned model directly on previously published data by comparing hand drawn histomorphometry outlines of the synovial tissue on slides (n=9) in the Test set from the TNF-Tg cohort (Batch A) published in Bell et al 2019 [18]. There was a significant positive correlation between the DL segmentation area and the hand drawn area (r^2^=0.96, **Figure 2A**) which demonstrates the accuracy of our method with hand drawn histomorphometry. 2) We then validated our model in a real-world setting by collecting 171 slides (64 knees, 2-to-3 histologic levels per knee) from 6-to-8-month-old TNF-Tg mice treated with Anti-TNF therapy for 6 weeks and slides from TNF-Tg and WT naïve and placebo-treated mice used as controls [19, 47] (**Figure 2B**). Male TNF-Tg mice display a robust inflammatory arthritis with synovial hyperplasia and pannus invasion of the distal femur and femoral articular cartilage starting around 3 months and reaching end-stage at 8 months of age, and our model measures this robust response well (**Figure 2C**, WT vs TNF-Tg, p<0.0001). Anti-TNF therapy is known to reduce synovitis yet does not alter trabecular bone loss in mice with established disease (>6 months old) [48, 49]. Our DL segmentation appropriately modeled these well-established structural changes autonomously (**Figure 2 C&D**), and it uncovered that cartilage degradation is also moderately reduced when Anti-TNF therapy is provided at 8 months of age for 6 weeks, which is an expected but novel result. Interestingly, trabecular bone area is already decreased in 6-month-old TNF-Tg mice compared to WT counterparts while cartilage area is not, suggesting that trabecular bone loss occurs before cartilage loss. Additionally, multiple other tissue structures can be studied simultaneously (**Supplemental Figure 7**) showing the versatility of studying H&E segmentation models to assess tissue structural changes within the context of mouse disease models. Taken together, these analyses suggests that our model accurately detects relevant and meaningful biologic treatment effects with the potential to discover novel structural changes. It is important to note that some of these slides were stained with a variation of traditional H&E, H&E-Orange-G (see *Methods*) demonstrating that our training strategy choices produced a model that is robust even when introducing stain variations.

**Figure 2.**
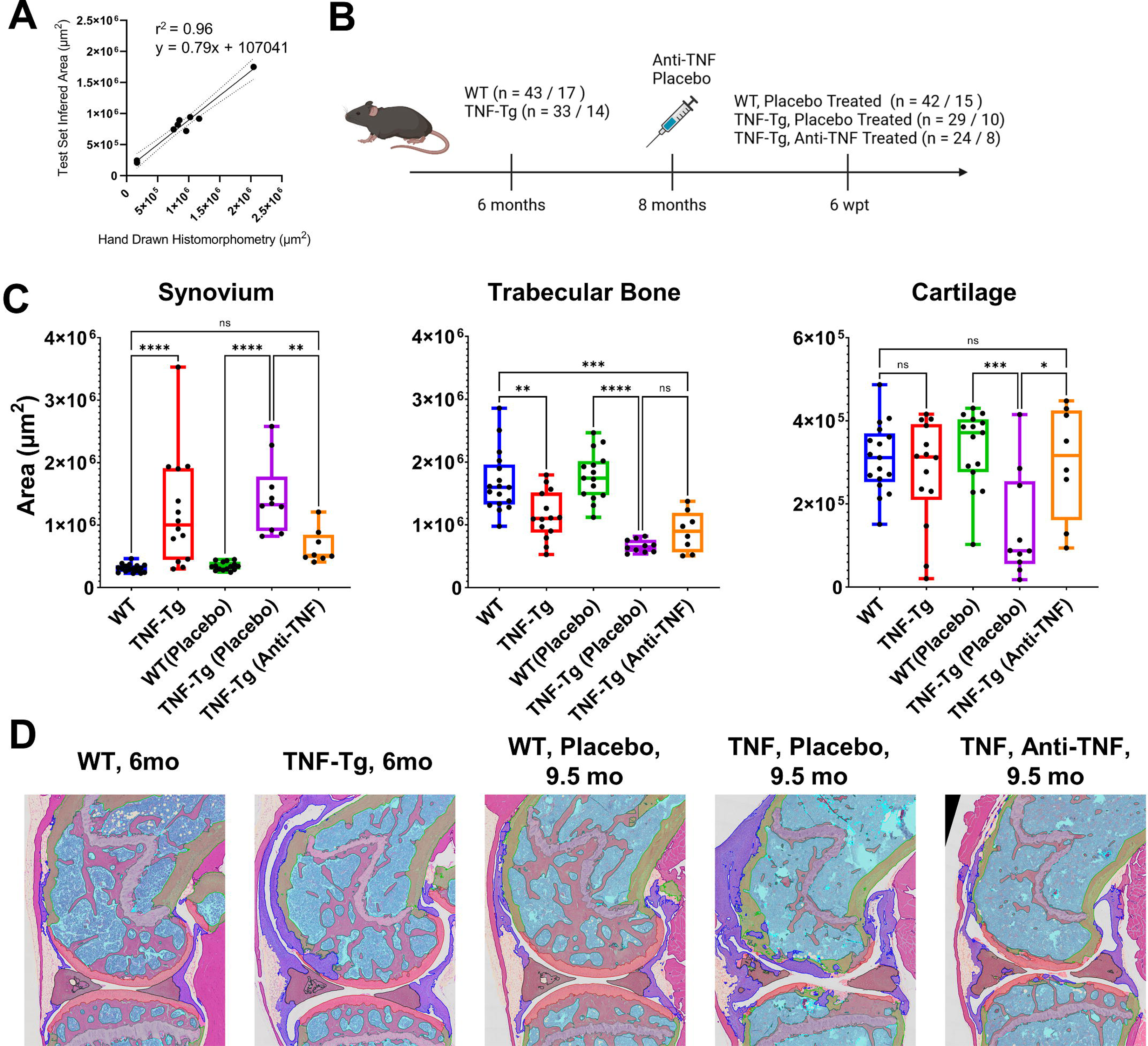
The fine tuned UNET++ model measures treatment response in the TNF-Tg with Anti-TNF therapy. Inferred test set synovial area correlated excellently with previously published hand drawn histomorphometry data (A). We then developed an independent cohort of slides from previously published work with a biologically relevant therapeutic intervention to test if our model is sensitive to detect these well described therapeutic response (B). As predicted from previous work, TNF-Tg have increased synovial area and lower trabecular bone area, with Anti-TNF therapy decreasing synovial area but does not alter trabecular area (C and D). Interestingly, we are able to detect the loss of cartilage area at end stage disease compared to younger TNF-Tg mice, and the therapeutic intervention protects from this loss (C, Right Panel).

### Tissue-Specific and Arthritis Effector Cell Types can be Identified with ML in TNF-Tg Mice

We annotated a total of 4,712 cells across three stages of murine inflammatory arthritis in the TNF-Tg mice (n=6 healthy, n=8 mild disease, and n=5 severe, **Supplemental Figure 8**) to build a cell type classification model that could recognize cells from various disease stages. We developed novel custom feature extraction methods, and the fidelity of these methods was demonstrated in 2D UMAP space, as most cell types are clearly separable (**Figure 3A**). We next built a GBDT classification model using stratified 5-fold cross validation training and tested our models’ predictions using three methods. First, we calculated the average (± SD of folds) F1 of each cell class in the test sets (**Figure 3B**). Each cell class demonstrated a good F1, between 0.61 for vessel cells and 0.92 for adipo-stromal cells with synovial associated cell classifications among the best (synovial lining cells = 0.87 ± 0.03; synovial fibroblasts = 0.79 ± 0.05; and synovial lymphocytes = 0.91 ± 0.05). To validate our classification, we used two strategies that utilized tissue and disease context. Tissue segmentation using the Original model was performed on these training slides and the remaining >300,000 cells cell-type classification was inferred. Tissues with known homogenous cell types, fat tissue and cartilage/meniscus, were investigated and cell types among these tissues were plotted. Within these adipose tissue and cartilage/meniscus, the most predicted cell type were adipo-stromal cells and chondrocytes, respectively (**Figure 3C**). Next, to assess predictions in the context of disease, we utilized the synovial tissue predictions only (as determined by the tissue segmentation model **Figure 1**) and stratified by disease severity. As shown by **Figure 3D**, synovial-specific increases in synovial fibroblasts, synovial lining cells and lymphocytes are seen with increasing disease severity. These results suggest that our cell type model can produce high quality predictions that are sensitive to disease stage.

**Figure 3.**
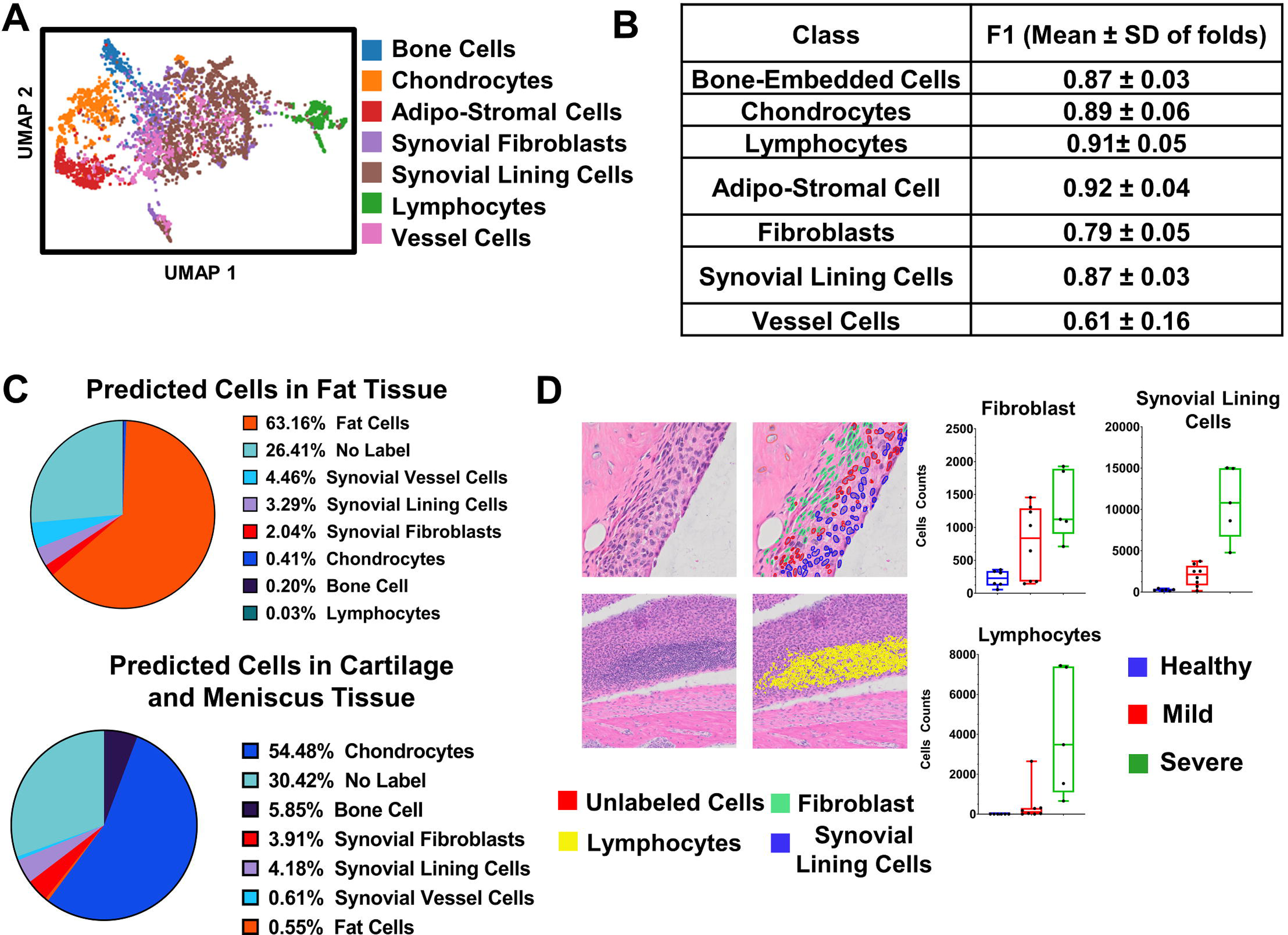
Cell type classification model successfully identifies important cell types in inflammatory arthritis. Utilizing Principal Component Analysis to reduce features and Uniform Manifold Approximation and Projection (UMAP) to project in 3D, we see good separation of Lymphocytes, Bone-Embedded Cells, Chondrocytes, and Adipo-stromal Cells (A). The gradient boosted decision tree performs well with an overall F1 of 0.83 ± 0.12 (M ± SD) and the class specific F1 is shown in (B). We then predicted the cell class (>75% predicted probability) of all other cells identified on the slides (∼300,000). To provide quality control, we looked at the predicted cells within the Fat tissue and Cartilage/Meniscus and our model appropriately classifies the majority cells within these tissue as fat cells or chondrocytes, respectively (C). We next looked at the cells within the synovium and found that our model classifies fibroblast, lymphocytes and other synovial lining cells well, and their distribution meets the expectation in healthy mild and severely disease knees (D).

Given the promising intra-test set performance and tissue- and disease-state specificity of the cell type modeling, we aimed to further validate our model with a larger data-set with more biologic variation. To do this, we utilize the previously described sexually dimorphic synovial pathology in TNF-Tg mice [18] and collected slides from 3-to-5.5-month-old WT and TNF-Tg-male and female mice. Confirming our previous observation using traditional histologic scoring, we found a significant increase of lymphocytes in female TNF-Tg synovium at 3 months of age with concomitant significant increase in synovial lining cells (**Figure 4A** and **B**).

**Figure 4.**
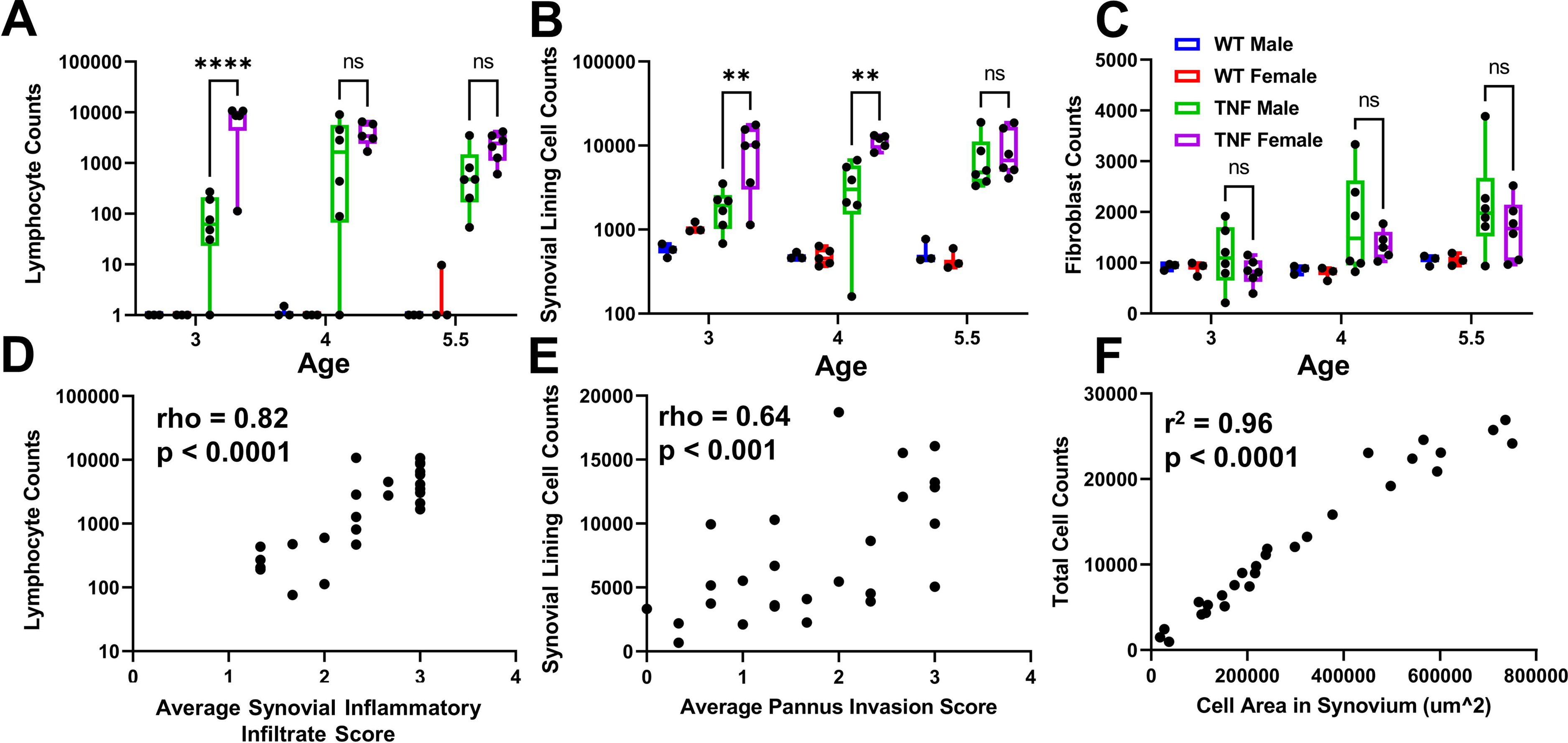
Cell type modeling recapitulates the sexual dimorphism of TNF-Tg inflammatory arthritis. Using this model to perform cell type predictions in a sexually dimorphic model of inflammatory arthritis, the computational pathology model describes previously demonstrated difference in inflammatory lymphoid infiltrates (here as Lymphocytes, **A**) and synovial hyperplasia (**B**, Synovial Lining cells). Interestingly, there is no difference in fibroblast cell (**C**) counts between TNF-Tg male and female mice. Importantly, we found a significant correlation between lymphocyte counts and the inflammatory infiltrate score (**D**); synovial lining cell counts and pannus invasion score (**E**); and between the total cell count and total cell area (as originally measured, **F**).

Interestingly, sexual dimorphism was not observed when assessing synovial fibroblasts, which is a novel finding (**Figure 4C**). Finally, we correlated our computationally derived lymphocyte and synovial lining cell counts with the synovial inflammatory score and pannus score, respectively (**Figure 4D** and **E**), and found good correlation between the parameters. This relationship also held for total cell counts (**Figure 4F**).

To reduce the annotation time, we explored various active learning approaches retrospectively in the murine cell type data set. Model performance using all the active learning strategies with 45% of the total training size was comparable to using the complete dataset with random sampling (0.8188 (0.8157-0.8219) vs 0.8213 (0.8184-0.8243), F1 ± SD). Additionally, with 45% of the data, the model performance using active learning was higher than using a randomly sampled set of examples (0.8188 (0.8157-0.8219) vs 0.8082(0.8053-0.8111) F1 ± SD) (**Supplemental Figure 10**). Additionally, the mean 5-fold CV macro F1-score was most different at the 10%-25% of annotated data range, indicating that active learning can drastically improve model performance with fewer examples.

### Cell Type Modeling on Human RA Synovial Biopsies Predicts Pathotypes and Correlates with Clinical Outcomes

After validating the active learning strategy in murine tissue, we next applied this approach to generate cell type annotations on human synovial tissue sections, aiming to reduce the overall histopathological evaluation time for our pathologist. We collected a small subset of initial annotations, predicted cell types on new cells and calculated the entropy-based uncertainty, ranking the most uncertain cells for future annotation by the pathologist. After multiple rounds with active learning, a total of 2,341 cells were annotated (**Supplemental Figure 11**). Using a gradient boosted decision tree with 5-fold cross validation we achieved model performance ranging from 0.74-0.88 F1-scores for all classes (**Figure 5A**). An example confusion matrix from the best performing fold demonstrates that undifferentiated stromal-connective cells are confused with vascular endothelial cells at the highest frequency and the plasma cells and lymphocytes are also often confused with each other (**Figure 5B**). Example prediction overlays demonstrate the high quality that our model produces for all our given cell types (**Figure 5C**).

**Figure 5.**
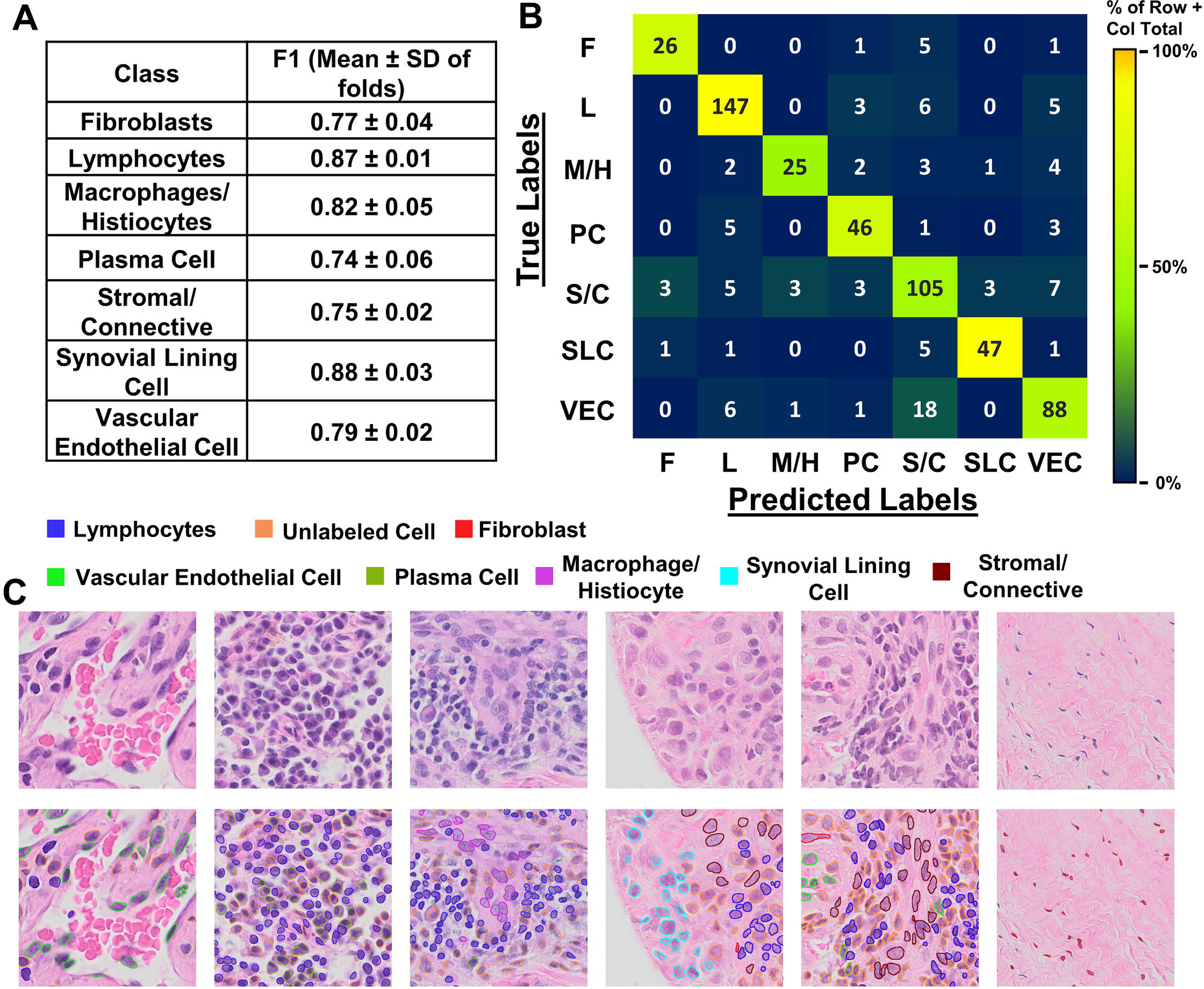
Cell type modeling correctly classifies synovial stromal and immune cells in RA synovial biopsies. The overall average F1 from the five-folds was 0.81±0.06, with all individual cell types performing well with greater than 0.75 F1 scores (A). Interrogating the confusion matrix (B) from the best performing model, we see that the most common mistakes are that plasma cell and lymphocytes are mistaken for each other and all the stromal cell populations (Stromal/Connective cells, Vascular Endothelial cells and Synovial Lining cells) can be confused. Representative images of cell class predictions with the original H&E on the top row and the cell type predictions overlays on the bottom row (C). F : Fibroblast, L : Lymphoid, M/H : Macrophage/Histocyte, PC : Plasma Cell, S/C : Stromal/Connective Cell, SLC : Synovial Lining Cell; VEC : Vascular Endothelial Cell.

To validate model performance, cell types were quantified for the entire synovial biopsy dataset with known across the three known pathotypes. Importantly, our cell type prediction a consistent with the previously described cellular distribution within each pathotype [10, 11, 14, 50]. Specifically, synovial fibroblasts are primarily found in the pauci-immune pathotype and some diffuse cases; while lymphocytes and plasma cells are found primarily in the lymphoid pathotype (**Figure 6A**). Interestingly, if a simple threshold is placed at 1.1% plasma cell to classify lymphoid (≥1.1%) or diffuse (<1.1%) cases, 19 out of 24 lymphoid cases and 22 out of 29 diffuse cases are correctly identified. Further, lymphocyte counts strongly correlates with the Krenn inflammation score (Spearman’s rho = 0.88, **Figure 6B**). Example images of the three pathotypes and their respective predictions demonstrate the cellular distributions of these pathotypes (**Figure 6C**).

**Figure 6.**
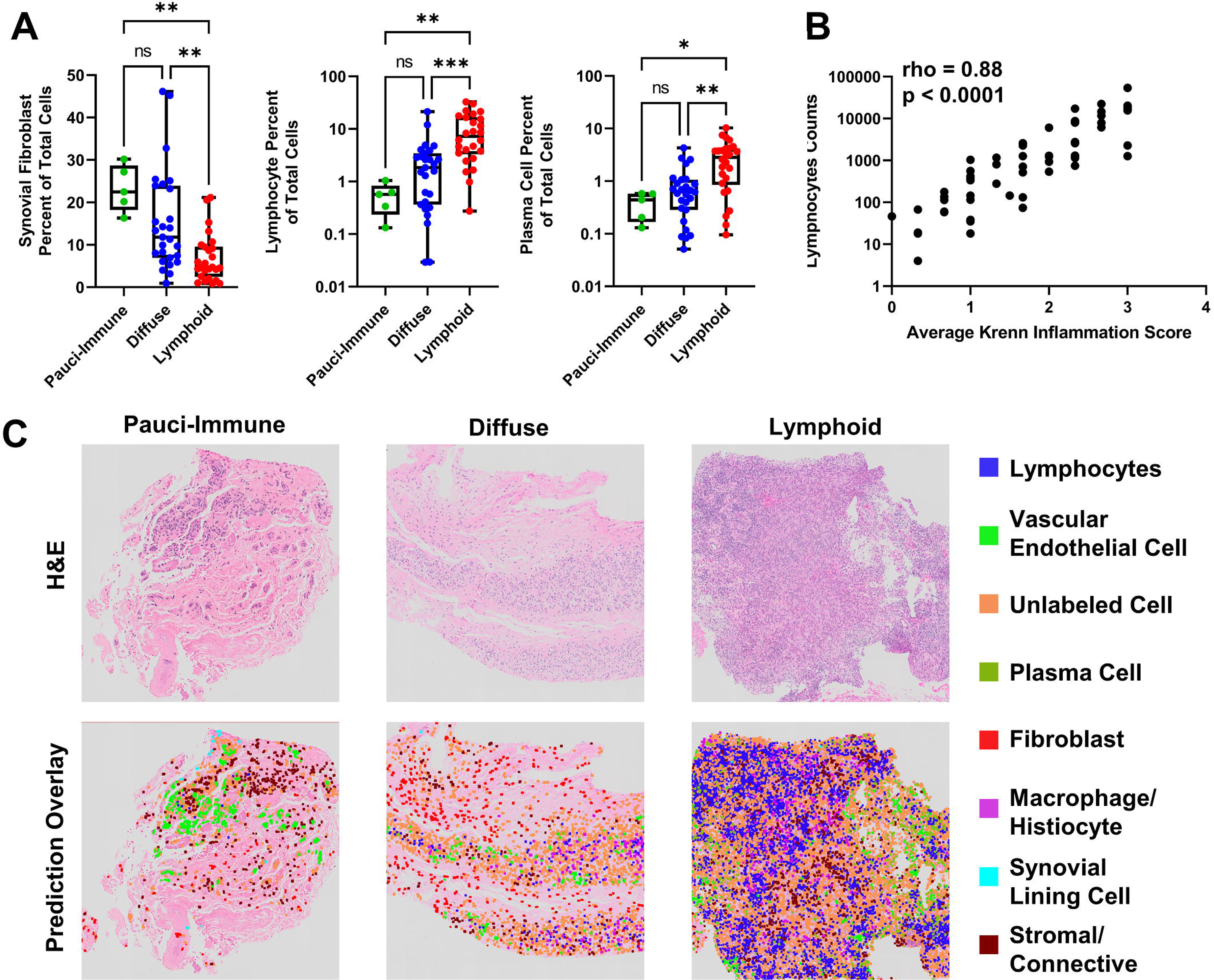
Cell type modeling associates with clinical pathotypes and correlates with clinical outcomes measures. Cell type predictions were made on 60 RA biopsy specimens (n=5 Pauci-Immune, n=27 Diffuse, n=26 Lymphoid) and plots of the Synovial Fibroblasts, Lymphocytes, and Plasma Cells percent of total cells demonstrate the known clinical differences among these pathotypes (**A**). We found a significant correlation between lymphocyte counts and the Krenn Inflammation score (**B**). Representative H&E and prediction overlay images (**D**) demonstrate the cellular diversity between the Pauci-immune, Diffuse, and Lymphoid pathotypes of RA.

## Discussion

Here, we demonstrate that multi-scale modeling of synovial histopathology can pathotype RA and inflammatory arthritis in clinically meaningful settings, such as treatment response. Our model will reduce the analytical bottleneck associated with histopathology assessment in both the clinical and preclinical settings allowing quicker times to intervention or hypothesis resolution. Furthermore, it will reduce the amount of accessory immunostaining required for pathotyping by using H&E stains to infer cell types, like lymphocytes and plasma cells, which would otherwise need an immunohistochemical (IHC) stain to confirm specific cell types. This could be very impactful as the diffuse pathotype (plasma cell poor with enrichment in myeloid cells) show improved response to tocilizumab (anti-IL-6R antibody) [50] while the fibroblast rich pauci-immune pathotype shows inadequate response to anti-TNF therapy [11]. While we did not have enough specimens to build a model to classify the pauci-immune pathotype in the AMP-RA data set, we were able to classify a diffuse vs lymphoid pathotype utilizing a simple threshold on plasma cell percent with a sensitivity and specificity of 0.81 and 0.73, respectively.

Computational approaches to understanding tissue and cellular information from histology slides have greatly improved in recent years. Specifically, tools to segment or classify of malignant tissues and cells from biopsy specimens [51], and cell type classification on cytology blood smears [52] have seen the largest amount of development with many applications acquiring FDA approval [53]. These tools have dramatically increase the throughput of clinical histologic analysis by, for example, red flagging potential malignant cases with >0.95 AUC for further review [54], classifying and counting cells on cytology smears or slides for detection of malignant cells and other pathologies [37, 52]. However, there is a dearth of computational pathology tools outside of cancer or outside of diagnostic settings and within the space of musculoskeletal pathologies [55]. Our work represents the first set of comprehensive tissue and cellular analysis tools for both pre-clinical and clinical phenotyping (in this case pathotyping) in inflammatory arthritis. In adjacent work, Pati and colleagues utilized hierarchical graph convolutional neural networks to integrate information from both the cellular and tissue levels on 2048px x 1536px sized images (0.42 uM^2^ pixels) [56]. This model was largely effective at detecting cancerous images with an F1 of 84.9% ± 0.8%, however did not perform well for non-cancerous, pre-cancerous or normal images with F1s of 56.6 ± 1.7%, 66.1 ± 3.7%, and 66.2 ± 1.7%, respectively. In other work, HoVer-Net is a state-of-the-art nucleus segmentation and cell type classification deep learning model for H&E-stained tissues [37]. However, the model was originally trained to identify malignant cells from other stromal (epithelium, fibroblast, muscle or endothelium) or inflammatory cells and only on small image patches. In synovial biopsies from RA patients’ further delineation of inflammatory cells is critical to pathotyping. Thus, we utilized the nuclear segmentation portion of the model and transferred this knowledge into our custom feature extraction pipeline to build a model to classify discrete immune cells relevant in RA synovial pathology.

Some computational approaches have already been successfully applied to H&E slides of synovial biopsies of RA patients to quantify cellular changes, such as nuclei density and its association with clinical inflammatory measures [57], and simple counting of CD3+ T-cells or CD68+ macrophages on IHC [58–60]. Further, pathologist scores of specific cells types have been associated with quantitative inflammatory gene expression changes in the RA synovium [61]. Our approach aimed to incorporate the important cell types from this previous work while producing models that only require H&E-stained tissues. However, RA has been shown to be a complex polygenic autoimmune disorder with various environmental risk factors contributing to multiple etiologies [12, 62, 63]. As a consequence of this complexity, many RA patients are refractory to existing approved therapies [64–67]. This highlights the need for a personalized medicine approach to improve clinical trial design and treatment allocation, as done in recent trials like the R4RA [6] and PEAC [68] that utilize ultrasound guided synovial biopsies and pathotype evaluation to stratify patients. Our models may be able to improve the workflow of these types of clinical trials and reduce the overall cost.

One major benefit of our model is the fact that we used H&E-stained tissues. These tissues are routinely and easily collected and represent a large proportion of historical datasets allowing for larger retrospective studies. While some of the computational pathology tools utilize H&E-stained slides, many have utilized special stains geared towards tissue- or cell type-specific classification [55]. This limits the overall throughput and utility of such pipelines by adding additional steps and costs that may be prohibitive. In addition, using a common stain facilitates transfer learning applications to other musculoskeletal pathologies, like osteoarthritis, bone fracture or disc degeneration, with different etiologies but similar tissue involvement and histopathology requirements.

Histopathology scoring that utilize Likert scales [69] is the gold standard analysis method of histology. For RA, various types of assessment, including Krenn lining and inflammation scores, rely on a consensus grading system to account for the challenge in analyzing H&E-stained tissue [70]. However, many scoring mechanisms quantify high level pathologic features (e.g. percent of area effected) or composite measures, without the level of detail included in our pipeline. The added benefit of quantifying cellular abundance and discrete tissue margins is that it can increase our ability to detect and predict the sensitivity to biological or therapeutic differences over Likert scale-based histology scores. For example, in our previous work we used a Likert scale to quantify synovial inflammatory infiltrate in which a score of 3 corresponded to “>30 inflammatory cells thick” while in our current work we use our computational approaches to count the specific number of lymphocytes that are present in the synovium. This is evident when the effect size (Glasse’s dela) is calculated for Female vs Male, TNF-Tg, 3-month-old of the scores vs the cell counts is compared (Histologic Scoring: 2.79 vs Lymphocyte Counts: 74.82) suggesting our cell classification model is orders of magnitude more sensitive to biologic differences than histologic scoring. These differences in quantification method represents ∼25 fold increase in measured effect size at 3 months between female and male TNF-Tg mice (Figure 5A) using the computational approach, suggesting a greatly enhance the ability to detect very small differences in therapeutic or other interventions to modulate the pathology. In addition to more granular quantification of pathology these analyses are also more efficient. For example, to generate the annotations to build our segmentation model we spent 200+ hours drawing annotations on the 94 slides. However, to infer the tissue segment on the 174 slides in Figure 2 the model took ∼30 hours of hands-off compute time with only 2-3 hours of labor to visualize the results, representing a ∼120-fold increase in efficiency.

Lastly, utilizing computational tools that quantify multiple tissues and cell types improves the ability to find novel phenomena. For example, while our primary focus is on the synovial pathology in inflammatory arthritis, having a model that measures cartilage, meniscus, and bone pathology provides a more comprehensive picture of disease. This allows easier detection of off-target or unexpected therapeutic effects with a singular methodology. However, reliance solely on computational modeling maybe increase false positives and expert-level quality control is advised for high impact results.

In conclusion, we have developed a set of models that can characterize tissue and cellular pathology in pre-clinical and clinical inflammatory arthritis settings. These models can be leveraged to better understand disease mechanisms in pre-clinical models and be used in a precision medicine pipeline to improve patients’ health.

## Data Availability

All raw and processed data will be available upon acceptance.

## Acknowledgments

Accelerating Medicines Partnership Program: Rheumatoid Arthritis and Systemic Lupus Erythematosus (AMP RA/SLE) Network includes:

Jennifer Albrecht^1^, William Apruzzese^2^, Brendan F. Boyce^1^, David L. Boyle^3^, Michael B. Brenner^2^, S. Louis Bridges Jr.^10^, Christopher D. Buckley^4^, Jane H. Buckner^5^, Vivian P. Bykerk^1,3^, James Dolan^2^, Thomas M. Eisenhaure^6^, Andrew Filer^4^, Gary S. Firestein^3^, Chamith Y. Fonseka^2,6^, Ellen M. Gravallese^7^, Peter K. Gregersen^8^, Joel M. Guthridge^9^, Maria Gutierrez-Arcelus^2,6^, Nir Hacohen^6^, V. Michael Holers^7^, Laura B. Hughes^10^, Eddie A. James^5^, Judith A. James^9^, A. Helena Jonsson^2^, Josh Keegan^2^, Stephen Kelly^11^, James A. Lederer^2^, Yvonne C. Lee^12^, David J. Lieb^6^, Arthur M. Mandelin II^12^, Mandy J. McGeachy^13^, Michael A. McNamara^1,3^, Joseph R. Mears^2,6^, Nida Meednu^1^, Fumitaka Mizoguchi^2,14^, Larry Moreland^13^, Jennifer P. Nguyen^2^, Akiko Noma^6^, Chad Nusbaum^6^, Harris Perlman^12^, Javier Rangel-Moreno^1^, Christopher T. Ritchlin^1^, William H. Robinson^24^, Mina Rohani-Pichavant^24^, Cristina Rozo^3^, Karen Salomon-Escoto^7^, Jennifer Seifert^7^, Anupamaa Seshadri^2^, Kamil Slowikowski^2,6^, Danielle Sutherby^6^, Darren Tabechian^1^, Jason D. Turner^4^, Paul J. Utz^15^, Gerald F. M. Watts^2^, Kevin Wei^2^, Costantino Pitzalis^16^, Deepak A. Rao^2^, Michael B. Brenner^2^, Soumya Raychaudhuri^2^

1University of Rochester Medical Center, Rochester, NY, USA.

2Brigham and Women’s Hospital and Harvard Medical School, Boston, MA, USA.

3University of California, San Diego, La Jolla, CA, USA.

4University Hospitals Birmingham NHS Foundation Trust and University of Birmingham, Birmingham, UK.

5Benaroya Research Institute at Virginia Mason, Seattle, WA, USA.

6Broad Institute of MIT and Harvard, Cambridge, MA, USA.

7University of Massachusetts Medical School, Worcester, MA, USA.

8Feinstein Institute for Medical Research, Northwell Health, Manhasset, New York, NY, USA.

9Oklahoma Medical Research Foundation, Oklahoma City, OK, USA.

10University of Alabama at Birmingham, Birmingham, AL, USA.

11Barts Health NHS Trust, London, UK.

12Northwestern University Feinberg School of Medicine, Chicago, IL, USA.

13University of Pittsburgh School of Medicine, Pittsburgh, PA, USA.

14Graduate School of Medical and Dental Sciences, Tokyo Medical and Dental University, Tokyo, Japan.

15Stanford University School of Medicine, Palo Alto, CA, USA.

16Centre for Experimental Medicine & Rheumatology, William Harvey Research Institute, Queen Mary University of London; London, UK

## Supplemental Figure Legends

**Supplemental Figure 1. Training, Testing and Validation with batch effect study design.** A) Description of Batch A from the TNF-Tg cohort and Batch B from the ZIA cohort. B) Description of the two training strategies, Mixed Training and Single Batch training.

**Supplemental Figure 2. Tissue Hierarchy Annotation Strategy.** Description of the hierarchical tissue classification scheme and which tissues are among each of the classes. The overall frequency of each class is also reported on the right.

**Supplemental Figure 3. Examples of Image Augmentation.** The original image is provided in A with Gaussian and Speckle Noise (B), Drop Out (C), Rotation (D), Hue and Saturation with Drop Out(E), and Color Temperature adjustment (F).

**Supplemental Figure 4. Using highly overlapping tiles with a per pixel majority vote produces the best performing model.** mIOU and fwIOU statistics of the three tiling mechanisms are show in A (*data is also presented in in Figure 1). Representative prediction overlays of the no overlap tiling strategy which shows very poor performance, and the 50% overlap tiling strategy which has a prediction artifact caused by poor predictions at the edges of the tiles (Arrows, B).

Supplemental Figure 5. A mixed training strategy with high augmentation builds a high performing segmentation model. Staining batch effect is a major issue with histology and we intentionally collected images in two different batches despite both being stained with H&E. The difference in RGB distributions between the two batches (A) is shown in 3D RGB space. The mean and frequency weighted IOU scores of all the mixed training models are similar and outperform the single batch training (B). During single batch training, high augmentation preform that best. The SLIC-RF model is provided as reference and underperforms compared to all DL models. Representative images of both batches with the H&E, GT, RF, No Augmentation and High Augmentation overlays are presented in C. Single batch high augmentation model qualitatively produces more segmentation artifacts (arrows).

**Supplemental Figure 6. Tables of class specific mIOU’s across all models.** The 7 class, 9 class, 10 class and 11 class model performance with each class is presented (A-C).

**Supplemental Figure 7. The UNET++ Fine Tuned 10 class model phenotypes the TNF-Tg with Anti-TNF therapy.** The remaining 5 tissue class predictions do not show any meaningful differences between the Anit-TNF and control samples.

**Supplemental Figure 8. Mouse Cell Type Classification scheme.** We selected 7 basic cell types to classify and annotated on healthy (n=6), mild disease (n = 8) and severely diseased (n=5) TNF-Tg slides. We annotated bone-embedded cells (A, n=312), vessel cells (B, n = 378), adipo-stromal cells (C, n = 506), fibroblasts (healthy, D; diseased, E; n = 749), chondrocytes (articular, F; growth plate, G; n = 625), lymphocytes (H, n = 467), and all other synovial lining cells (healthy, I; diseased, J; n = 1675).

**Supplemental Figure 9. Active learning can reduce the number of training samples.** Three independent active learning statistics were evaluated (Entropy, Blue; Margin, Red: Uncertainty, Purple) to select cells and compared against randomly selecting cells (Green). Using the any of the active learning uncertainty calculations to select new training data to add outperforms random sampling overall (A, green curve is lower between 5-60%). Bone-embedded cells, Synovial Fibroblasts, synovial fats cells and synovial vessel cells all demonstrate improved performance with active learning while lymphocytes, synovial lining cells and growth plate chondrocytes do not (B-H). Interestingly, maximal training occurred with only ∼30% of the total training data set, suggesting this mechanism could be used to generate new annotations on a novel dataset. Note the different scale on D.

**Supplemental Figure 10. Human Cell type Classification scheme for Synovial Biopsy Specimens.** We identified 13 cases from the Accelerating Medicine Partnership – Rheumatoid Arthritis (AMP-RA) Phase 2 cohort that were distributed between the three RA synovial pathotypes (Lymphoid, n = 5; Diffuse, n = 5; Pauci-Immune, n = 3). A board-certified pathologist annotated 7 cell types among these 13 cases: stromal/connective tissue cells (A, n=541), synovial lining cells (B, n= 314), fibroblasts (C, n = 180), vascular endothelial cells (D, n = 349), Tissue Macrophages/Histocytes (E, n = 176), lymphocytes (F, Green Arrows; n = 555), and plasma cells (F, Blue Arrows; n = 226). In order to obtain these annotations, we used the uncertainty sampling active learning strategy as presented in Supp Figure 9 to select cells for our pathologist to annotate. The table describes the general hierarchy of cell labels, for example, if the pathologist could not further distinguish a stromal cell to be either a vascular endothelial cell or fibroblast, then the cell would be labeled as a stromal/connective cell.

