## Supplementary figures and images for "Automated multi-scale computational pathotyping (AMSCP) of inflamed synovial tissue"

### Supplemental Figure 1

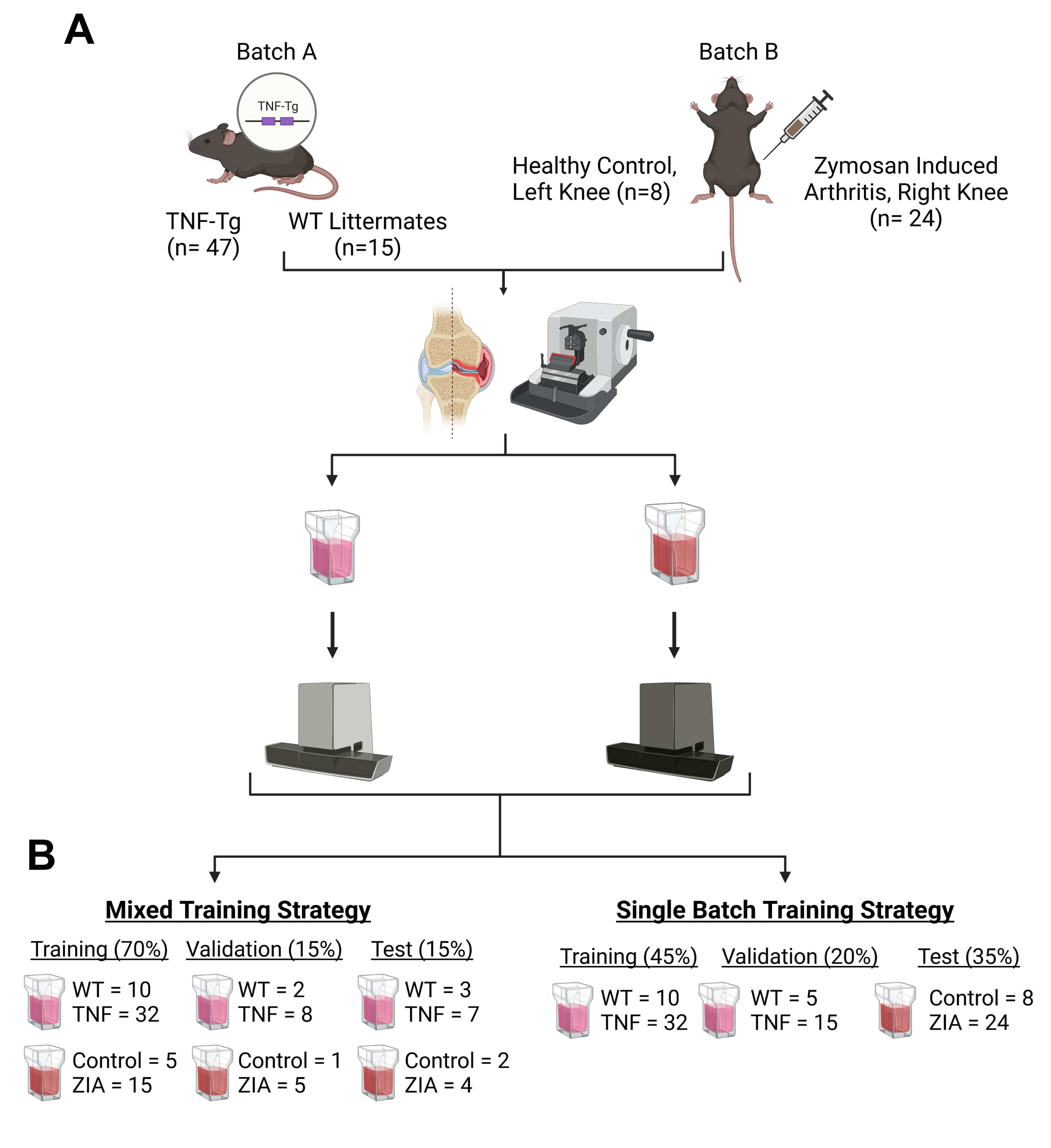

### Supplemental Figure 2

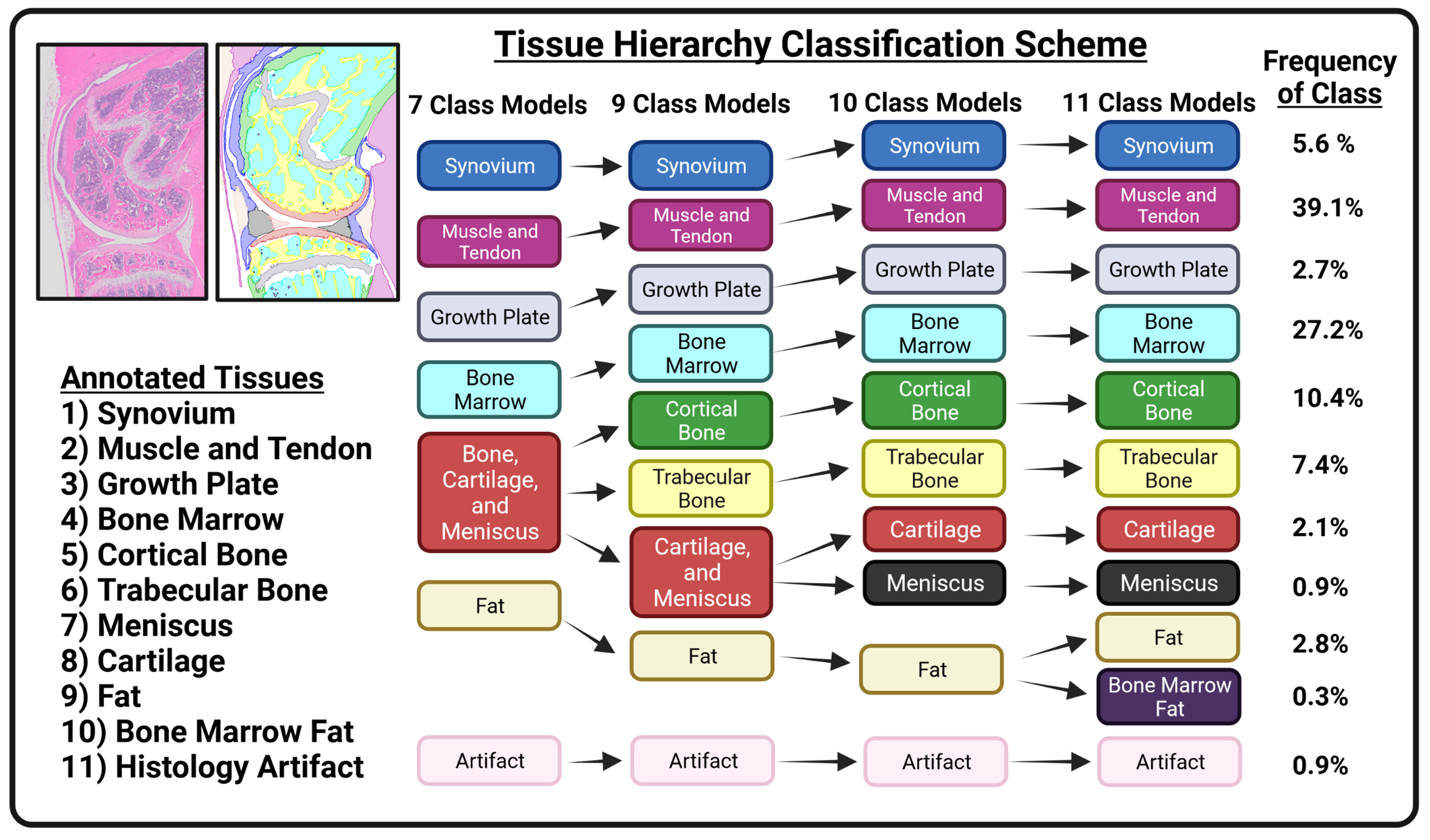

### Supplemental Figure 3

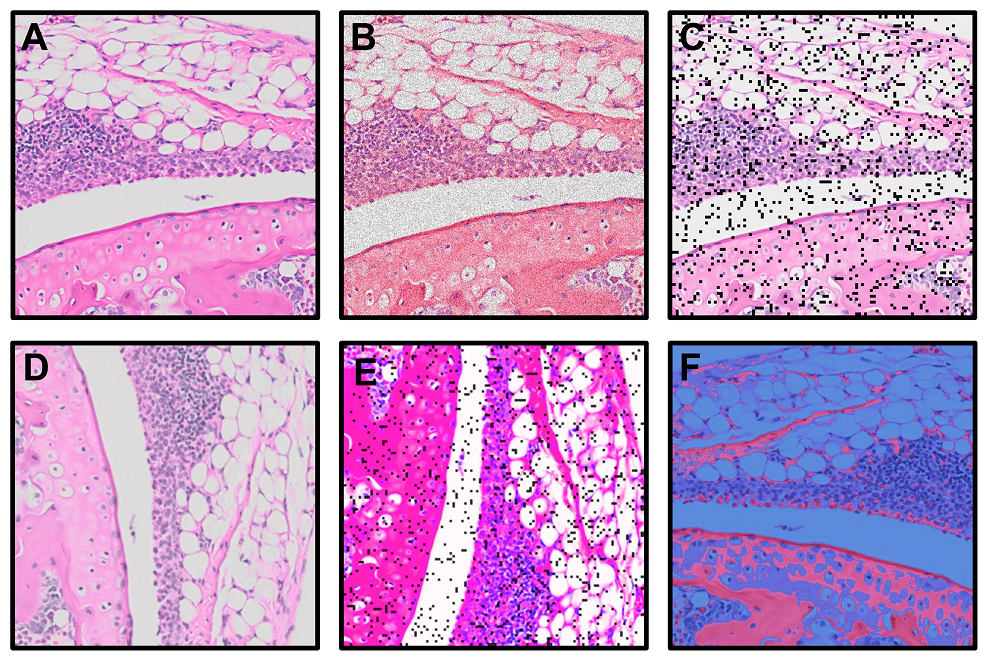

### Supplemental Figure 4

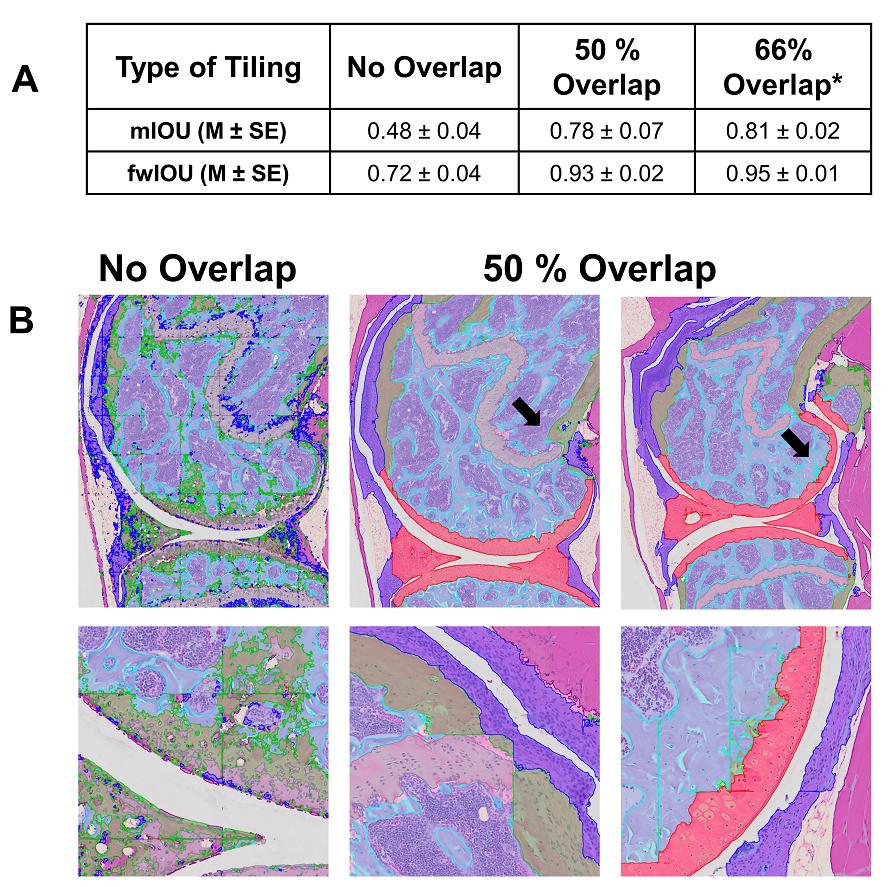

### Supplemental Figure 5

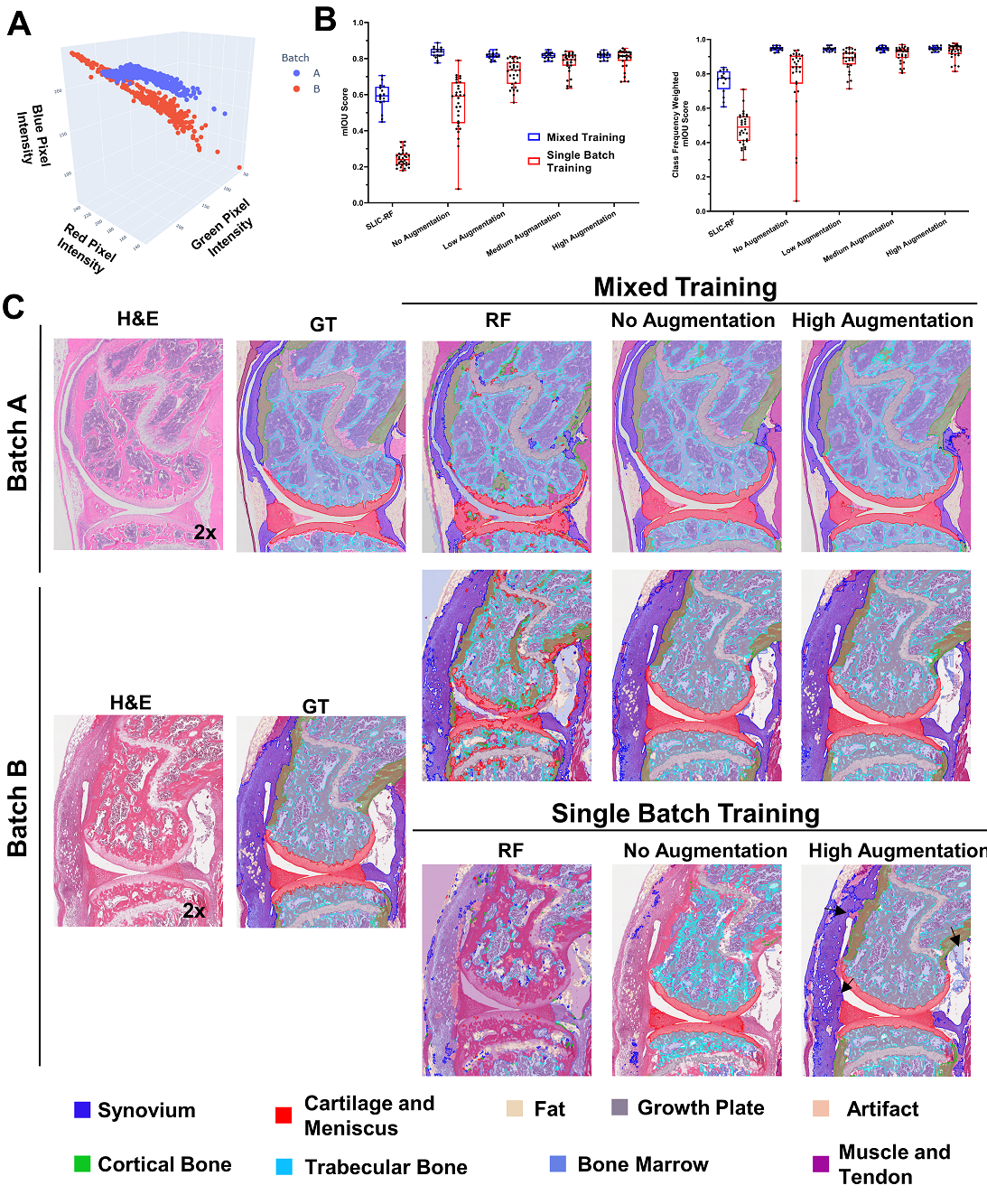

### Supplemental Figure 6

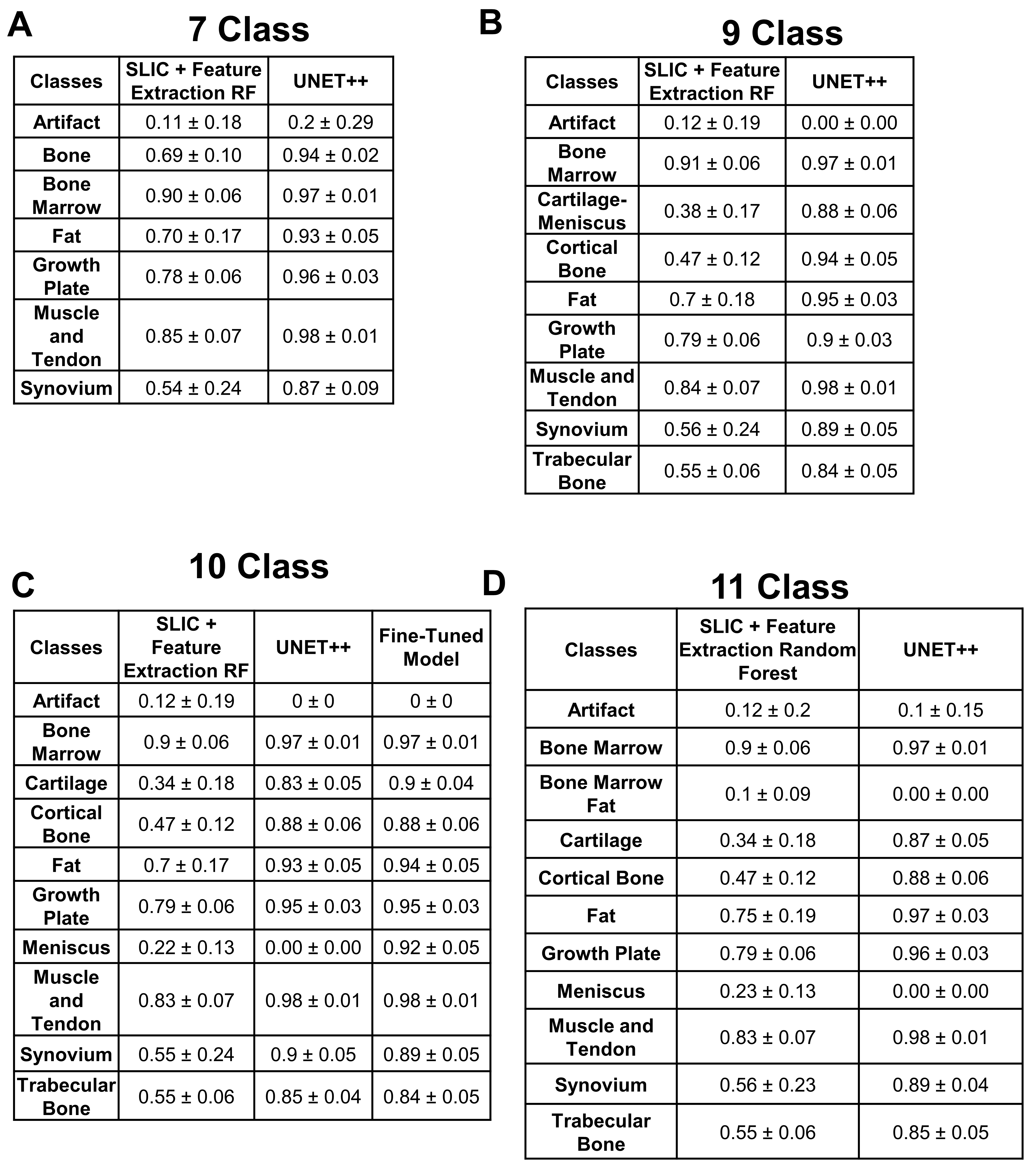

### Supplemental FIgure 7

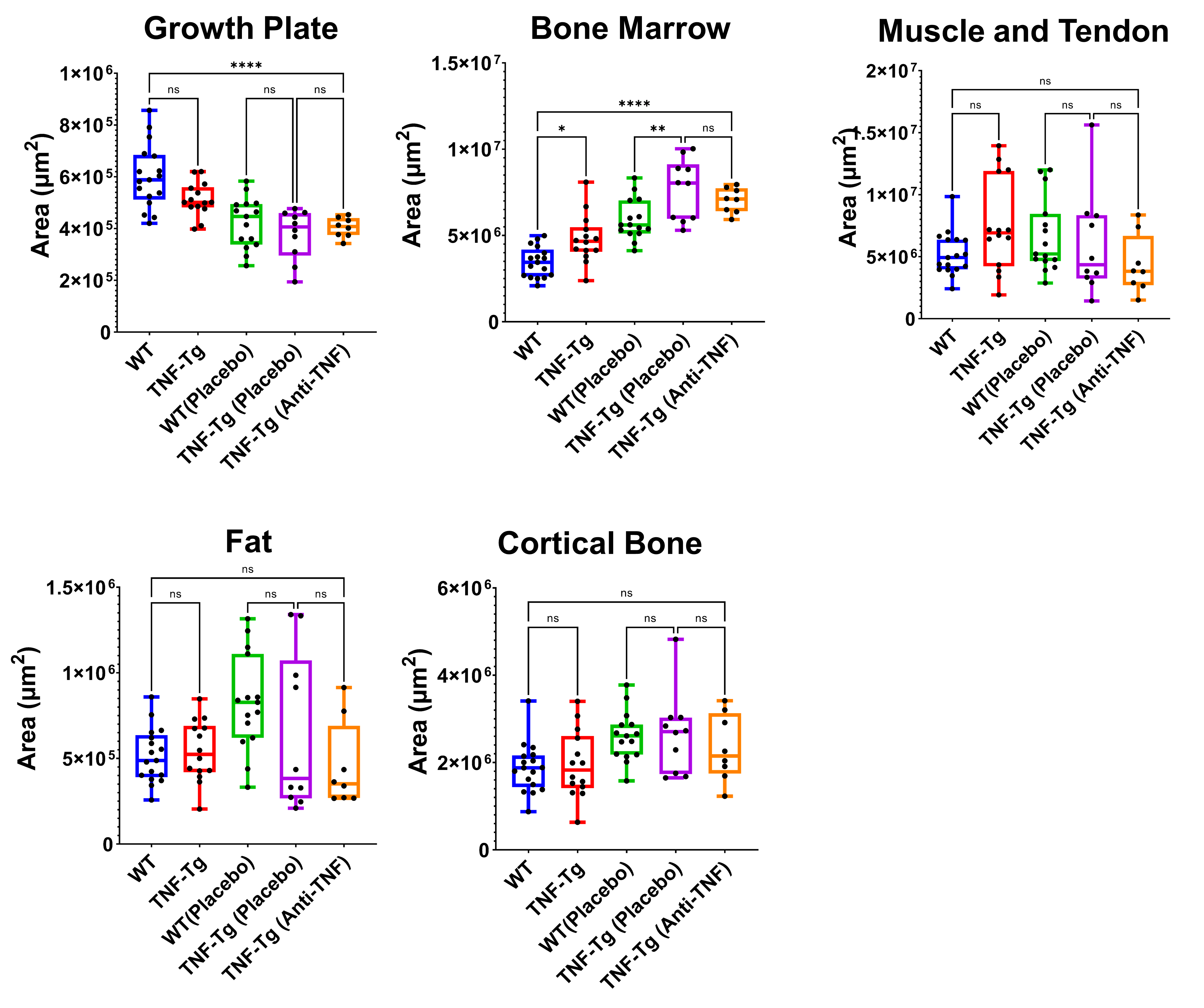

### Supplemental Figure 8

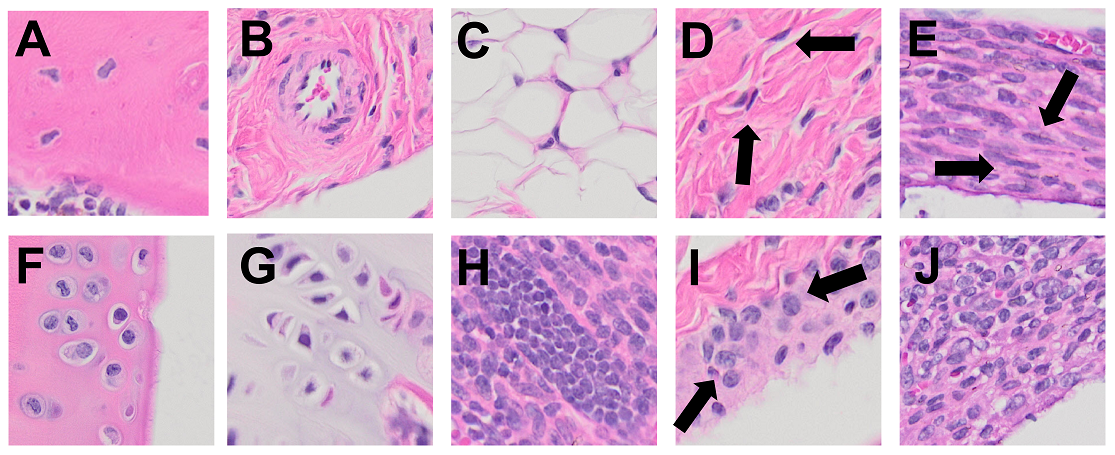

### Supplemental Figure 9

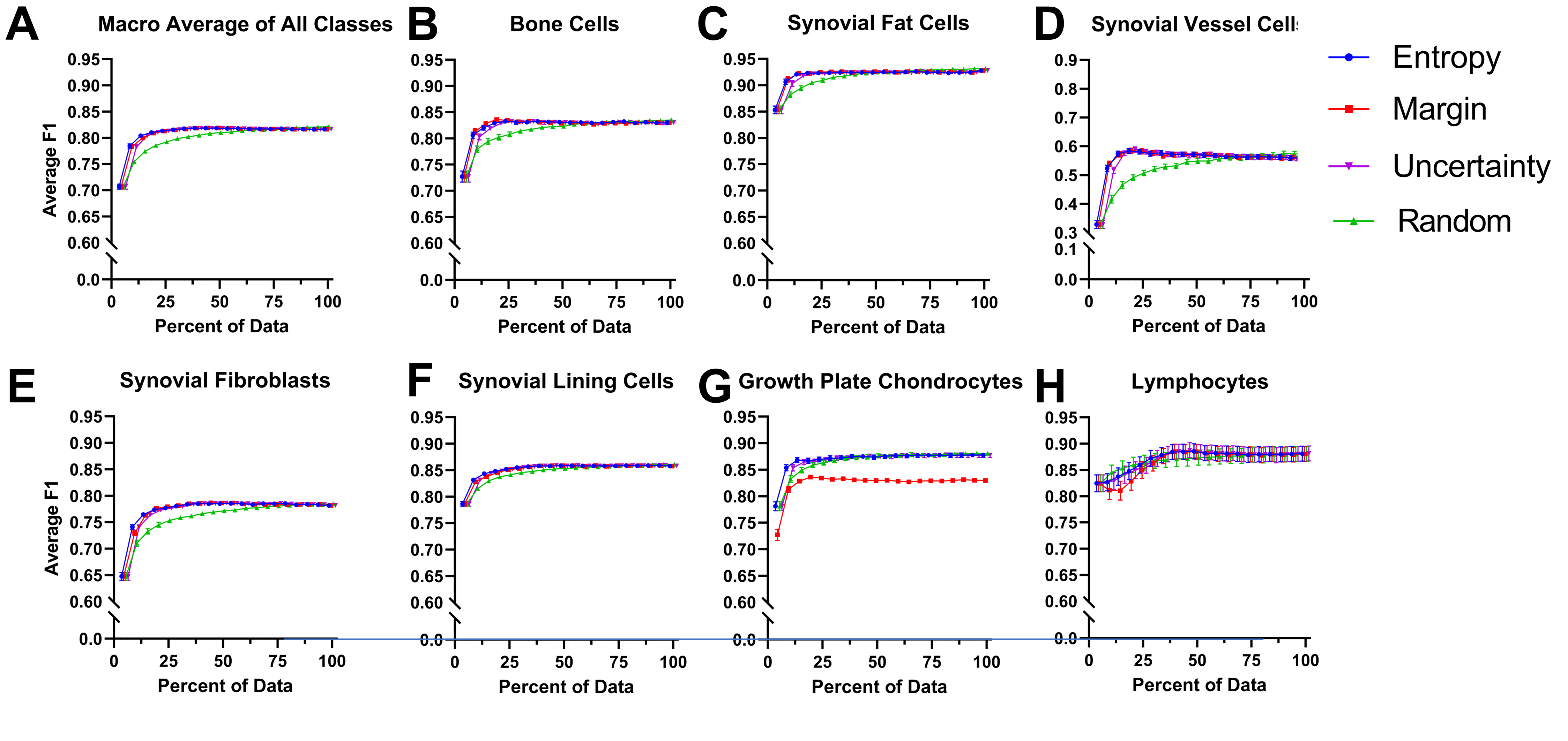

### Supplemental Figure 10

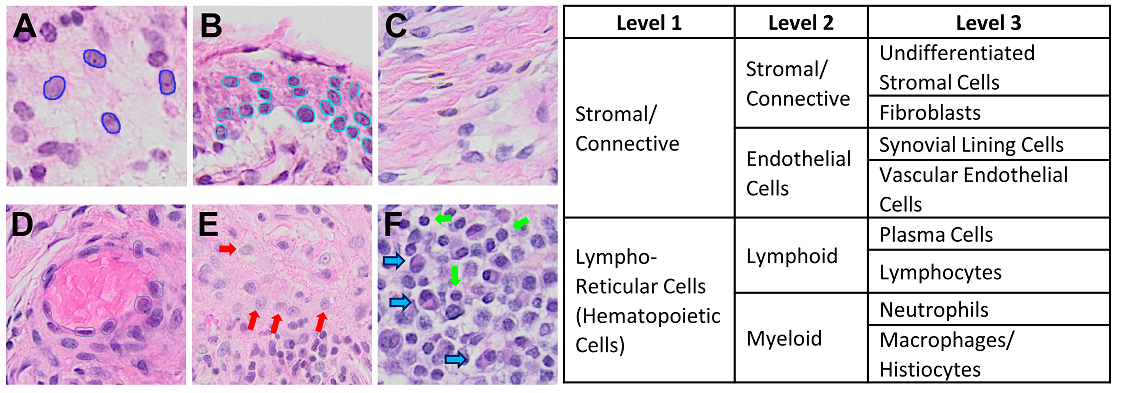
